# Impact of Transition from Conventional Open Radical Cystectomy to Laparoscopic Radical Cystectomy for Neobladder: A Retrospective Study

**DOI:** 10.1101/2022.03.20.22272324

**Authors:** Rupesh B Shah, Amit D Trivedi, Ketan B Rajyaguru, Parloop A Bhatt

## Abstract

**Background:** Early operative recovery and good Quality of life are important goals of radical cystectomy. We compare the pre, peri and post operative data between Open radical cystectomy (ORC) and Laparoscopic radical cystectomy (LRC) surgery of neobladder.

**Patients and Methods:** Retrospective analysis of 13 male consecutive patients who underwent radical cystectomy by a single surgeon was done. Diagnosis of all patients was of invasive bladder cancer. Abdominal and preoperative staging was done using computed tomography. None of them received neoadjuvant chemotherapy. All the patients received same standard template bilateral pelvic lympadenenectomy. The urinary diversion included orthotopic neobladder. All patients were consented prior to study participation.

**Results:** Of the 13 male patients, six had ORC with neobladder while 7 underwent LRC surgery. Baseline characteristics (age, BMI, comorbidities, tumour grade, lymph node status) were similar in both groups. Incision length was significantly smaller in LRC as compared to ORC group (p <0.0001). Although the operative time was longer in LRC group as compared to ORC it was sufficed by reduced time for analgesics, shorter hospital stay (p<0.05), besides earlier time to liquid intake with immediate removal of nasogastric tube (p<0.001). No major complications were observed in the LRC unlike ORC group where one patient died at 30 days.

**Conclusions:** Based on the observations of our small study sample peri and postoperative outcomes are promising for LRC compared to ORC for patients undergoing neobladder in terms of the smaller incision length associated with less pain and complications, with speedy recovery without jeopardizing oncological outcomes. Transition of surgeon from ORC to LRC was advantageous to patients.

## INTRODUCTION

Traditionally, Open Radical Cystectomy (ORC) is considered as the gold standard for treatment of localized muscle-invasive, bladder cancer^1^. Despite a better understanding of pelvic anatomy and improved surgical techniques, ORC is associated with significant perioperative complications like intraoperative blood loss even when performed by experienced surgeons^2-8^. With the advancement of laparoscopic equipment and skill of urologists, laparoscopic radical cystectomy (LRC)/Robotic assisted radical cystectomy (RARC) stands as an alternative to ORC^9-10^. LRC leads to faster recovery, reduced hospital stay and morbidity, prompt return to routine activities, maintaining functional and oncological outcomes similar to ORC^11-13^.

The prolonged LRC operative time is due to the urinary diversion rather than removal of the bladder^14^. To date, urinary diversion can be performed by 2 approaches i) intracorporeal and ii) extracorporeal approach for either ileal conduit or neobladder formation^14^. However, extracorporeal diversion is preferred since it helps reduce the operative time with comparable intracorporeal postoperative results^15^. A study reported neobladder formation 2.5 times more prevalent than conduit formation (130 conduits: 315 neobladders)^16^.

This manuscript reports the upgradation journey of surgical and scientific skills of the urology team in the last decade resulting in switch over from ORC to LRC; maneuvering from ileal conduit to neobladder diversion at a single center in a developing nation. This case series is a comparative analysis of ORC versus LRC of neobladder diversion in a similar set of patients establishing the superiority of LRC over ORC.

## PATIENTS AND METHODS

Total 55 radical cystectomies (RC) were performed at a tertiary referral hospital in Western India from June 2018 to January 2021. Forty-two who underwent ileal conduit (including women) were excluded from the study. We retrospectively analyzed 13 male consecutive patients who underwent radical cystectomy by a single surgeon. All patients had invasive bladder cancer. <u>A</u>bdominal and preoperative staging was done using <u>computed tomography</u> (CT). The possible benefits and drawbacks of LRC and ORC were explained and the patients could choose either open or laparoscopic surgery. Six patients choose traditional ORC while 7 opted for laparoscopic approach. None of them received neoadjuvant chemotherapy. All patients received the same standard bilateral pelvic lymphadenectomy template. The urinary diversion included the orthotopic <u>neobladder</u>. All patients gave a written consent for participation in the study.

### Surgical Technique

#### Laparoscopic Assisted Radical Cystectomy

Following general anesthesia, patient was placed in a low-lithotomy position. A five-port trans peritoneal approach was used. The first 10-mm camera trocar was placed at the <u>umbilicus</u> using an open technique. After a pneumoperitoneum was established, two 10-mm working ports were placed (right port kept above line between ASIS and umbilicus Para rectal near the midline, left port midway between anterior superior iliac spine (ASIS) and umbilicus). Two 5 mm port were kept just above ASIS on each side. Steep trendelenburg position was given, and bowel was reflected up. Both the ureters were isolated till the level of the bladder and Hem-o-lok clips were used for ligation. The bladder was then dissected from adjacent tissues and the bladder pedicles were divided with ligasure. Bladder was dropped down and anteriorly space of Retzius was entered, B/L pubo prostate ligament were divided, endopelvic fascia was incised and deep venous complex was ligated with a V lock (3-0). Urethra was transected and frozen section was sent as and when needed to avoid positive surgical (PSM) margin. B/L pelvic lymph node dissection was done. Through small (around 7-8 cm) lower midline incision specimen was delivered out, orthotopic ileal neobladder was prepared by Studer technique^17^. Energy source utilized was ligasure, bipolar cauterized as and when required.

### Open Radical Cystectomy

Open radical cystectomy was done through standard lower midline incision (20-22 cm) extending just above the umbilical and orthotopic ileal neobladder was prepared.

In both groups suprapubic catheter (SPC) used was 14 F while per urethral catheter (PUC) was 20 F. SPC and PUC were removed accordingly on 14^th^ and 21^st^ post-operative day (POD). DJ stent for ureteroileal anastomosis was placed, which was removed at 6 weeks.

### Pre –and Peri- operative Management

Preoperatively, all patients received liquid diet 24 hours prior to surgery. No mechanical or antibiotic bowel preparations were given to any patients. Prophylactic single-dose second-generation cephalosporin was given at induction along with metronidazole before opening of the bowel. If the duration of the procedure was greater than 4 hours, use of antibiotics was repeated. To none of the patient low molecular weight heparin was given prophylactically. Removal of nasogastric tube and oral liquid intake was initiated as per patient’s recovery. Prokinetic (lesuride) was given to enhance bowel mobility.

Patient characteristics including age, body mass index (BMI) and co-morbidities were assessed. Peri-operative measures were compared including estimated blood loss, operative time and transfusion needed. Hemoglobin (Hb) and serum creatinine (SCR) were investigated pre and post-surgery. Post-operative parameters included-time to liquid intake, time to removal of nasogastric tube, average length of hospital stay, days of use of analgesics and post-operative complications. Oncological parameters included tumor stage, lymph node status and lymph node yield. Survival analysis was performed at hospital discharge and 30 days’ post discharge.

### Statistical Analysis

Categorical variables are expressed as number and percentage, with comparison between groups using Pearson Chi square or Fisher’s exact tests. Continuous variables were expressed as mean and range with comparison between groups using Manne Whitney test. Statistical analysis was performed using SPSS version 19. The values are compared at 95% confidence interval (CI) where, p <0.05 was considered statistically significant difference.

## RESULTS

Of the total 13 RC male patients, six underwent ORC while seven underwent LRC with neobladder diversion. Table 1 depicts the patients’ demographic data. The mean age was 59.1 years in the LRC group and 53.3 years in the ORC group. There were no significant differences in age, BMI, Hb, SCR and co morbidities between the two groups.

**Table 1:** Baseline Characteristics.

| Characteristics | ORC group (n= | LRC group | p value | 95% CI |
| --- | --- | --- | --- | --- |

|  | **6)** | **(n= 7)** |  |  |
| --- | --- | --- | --- | --- |
| Age (years) | 53.3 (29-69) | 59.1 (52-65) | 0.3526 | -7.35% to -18.95% |
| BMI* (kg/m2) | 23.05 (20.6-25.6) | 24.18 (19.5-29.5) | 0.5422 | -2.82% to -5.08% |
| Hb<sup>†</sup> (g/L) | 10.6 (7.1-13.9) | 12.22 (10.8-14.9) | 0.1745 | -0.83% to -4.07% |
| SCR<sup>‡</sup> (umol/L) | 1.08 (0.67-1.37) | 1.15 (0.79-1.87) | 0.7304 | -0.36% to -0.50% |
| **Comorbid Conditions** |  |  |  |  |
| Hypertension | 1 (16.7) | 4 (57.14) | 0.1512 | -10.59% to 70.74% |
| Diabetes mellitus | 1(16.7) | 1 (14.28) | 0.9078 | -37.05% to 43.79% |
| Hypothyroidism | 0 | 1 (14.28) | 0.3546 | -26.47% to 51.30% |
| COPD | 0 | 0 | -- | -- |

*Data are presented as n, n (%) or mean range.*
*\*BMI= Body mass index, <sup>†</sup>Hb= Hemoglobin, <sup>‡</sup>SCR= Serum creatinine*

### Pathological Features

Table 2 presents the pathological features. Tumor stage, grade, lymph node status between the two groups were similar (p> 0.05). Number of lymph node dissected was comparably less in the LRC group (12.28 vs. 19.16).

**Table 2:** Pathological Features.

| Pathological factors | ORC group (n=6) | LRC group (n=7) | p value | 95% CI |
| --- | --- | --- | --- | --- |
| Tumor stage |  |  |  |  |
| T0, Ta, Tis,<br>T1 | 1(16.7) | 1 (14.28) | 0.9078 | -37.05% to 43.79% |
| T2 | 2 (33.3) | 3 (42.85) | 0.7346 | -36.01% to 49.41% |
| T3 | 2 (33.3) | 1 (14.28) | 0.4355 | -24.90% to 57.52% |
| T4 | 1(16.7) | 2 (28.57) | 0.6267 | -32.72% to 49.94% |
| **Lymph node status** |  |  |  |  |
| Negative | 4 (66.66) | 5 (71.42) | 0.8587 | -37.92% to 46.69% |
| Positive | 2 (33.3) | 2 (28.57) | 0.8595 | -37.94% to 46.67% |
| Dissected<br>Lymph node<br>number | 19.16 ± 2.48 | 12.28 ± 4.49 | 0.0067 | -11.42% to 2.33% |
| Positive<br>Surgical<br>Margin | 0 | 0 | - | - |

*Data are presented as n, n (%) or mean ± SD.*

### Operative and Postoperative Characteristics and Management

Table 3 depicts operative and postoperative characteristics. Incision length was significantly smaller in LRC patients as compared to ORC group (20.83 ± 0.75 vs 7.14 ± 0.37 cm, P <0.0001). Figure 1 and 2 are schematic representations of each group depicting incision length. Also blood loss in LRC group was significantly less as compared to ORC group (P <0.0001). Requirement of blood transfusion was nearly similar in either groups. However, operating time was significantly longer in the LRC group as compared to ORC group (384.16 ± 47.68 vs. 522.14 ± 45.63 min.). Hb and SCR levels were not affected by the type of surgery.

**Table 3.**
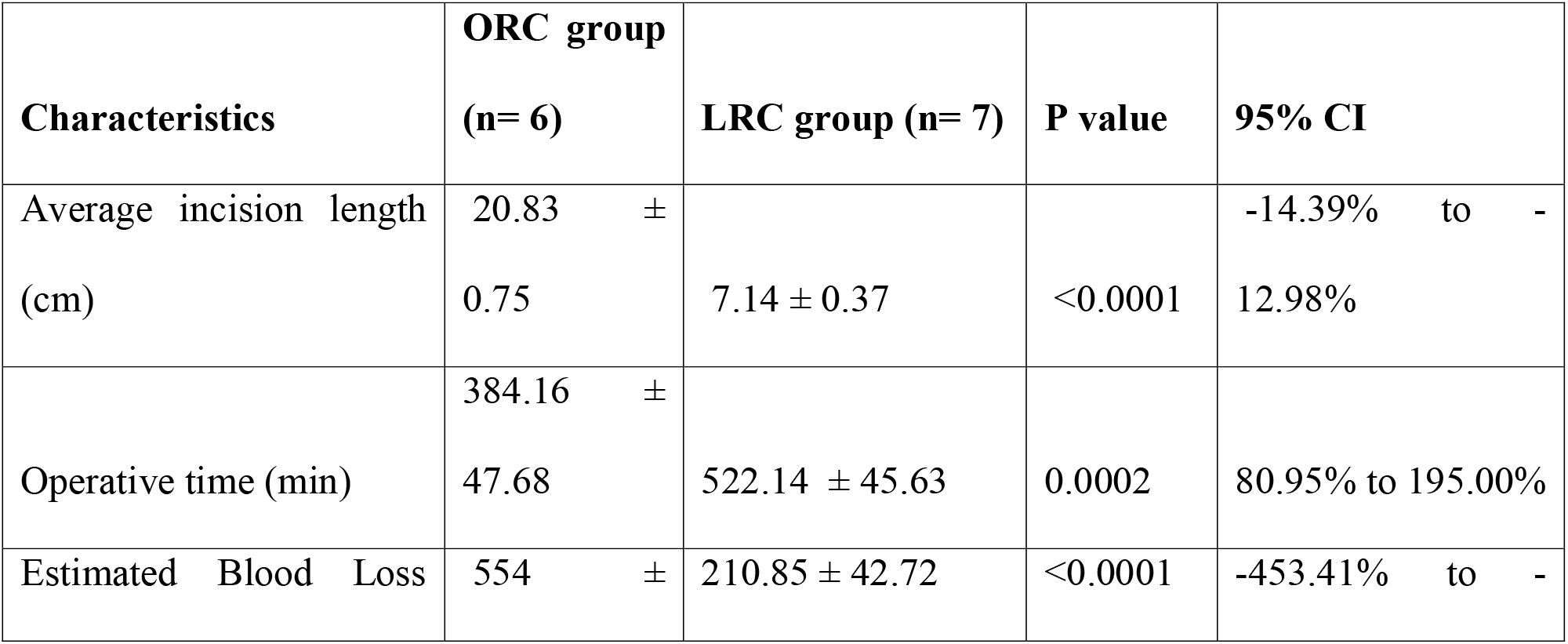
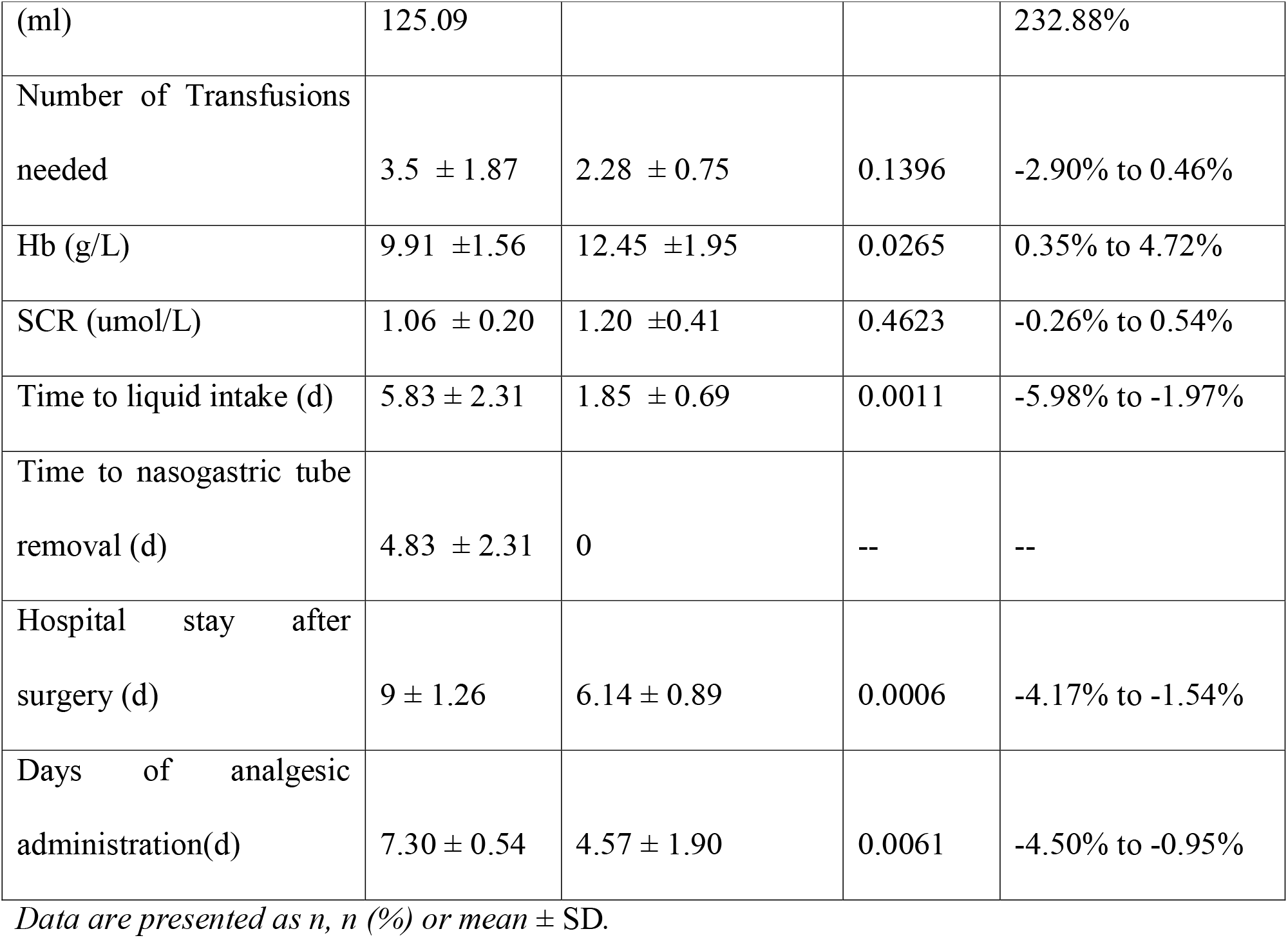
Operative and Postoperative Characteristics.

Patients in LRC group had earlier liquid intake as compared to ORC group. Naso-gastric tube was immediately removed post operatively in LRC group, while the removal of the tube in ORC group was at 4.83 days. Patients with LRC had significantly shorter hospital stay as compared to ORC patients (9 ± 1.26 vs. 6.14 ± 0.89 days). Patients undergoing LRC had pain for longer duration post operatively and thus were on analgesics for 7.30 ± 0.54 days as compared to ORC patients (4.57 ± 1.90 days). No positive surgical margins (PSM) were noted in any patients.

### Perioperative Complications

Table 4 reports the complications observed in both groups. A total of 8 complications were observed in the ORC patients as compared to LRC patients wherein only 1 patient reported atelectasis which was managed in ward with high flow oxygen and intensive physiotherapy using spirometer. The complications in the ORC group included-ileus in 5 patients, of which one patient underwent reoperation besides infection and wound dehiscence.

**Table 4.**
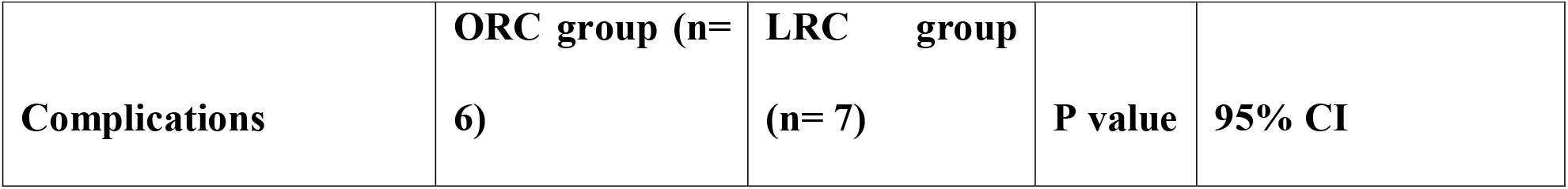
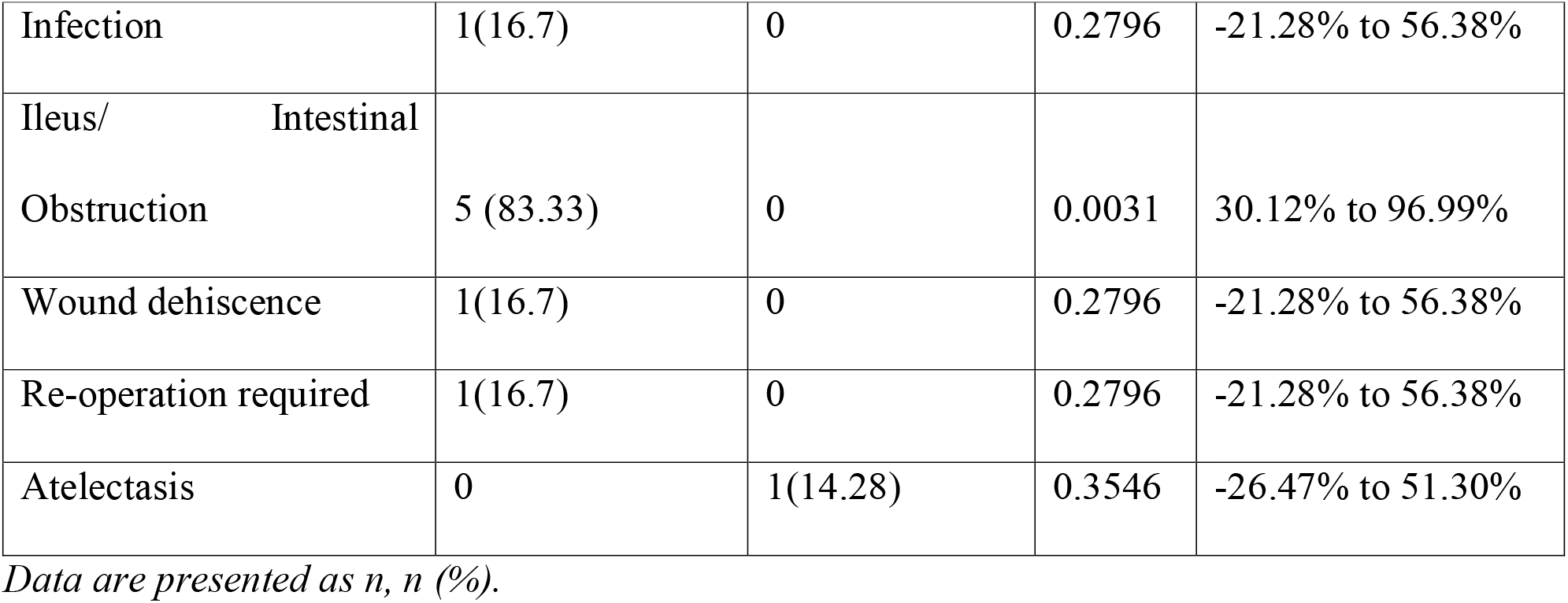
Postoperative complications of the two groups.

| Complications | ORC group (n= 6) | LRC group (n= 7) | P value | 95% CI |
| --- | --- | --- | --- | --- |
| Infection | 1(16.7) | 0 | 0.2796 | -21.28% to 56.38% |
| Ileus/ Intestinal<br>Obstruction | 5 (83.33) | 0 | 0.0031 | 30.12% to 96.99% |
| Wound dehiscence | 1(16.7) | 0 | 0.2796 | -21.28% to 56.38% |
| Re-operation required | 1(16.7) | 0 | 0.2796 | -21.28% to 56.38% |
| Atelectasis | 0 | 1(14.28) | 0.3546 | -26.47% to 51.30% |

*Data are presented as n, n (%).*

### Outcomes- At Hospital Discharge and 30 Days

No in-hospital mortality was reported in either groups. All patients in both groups were discharged in a hemodynamically stable condition. At 30 days follow up, no mortality was observed in LRC group. One patient in the ORC group, reported intestinal obstruction, abdominal burst and sepsis resulting to death at 30 days.

## DISCUSSION

Although not many studies of neobladder diversion are available comparing ORC and LRC, orthotopic neobladder has several advantages including voiding through natural passage, avoidance of external appliance, preservation of body image and superior quality of life^18^. Recently Yuan-hua Liu et al., showed LRC plus neobladder short term curative efficacy with early recovery and good neobladder function^19^. Robotic techniques are increasingly advocated because of their advantages, like reduced blood loss and analgesic requirements with quicker recovery^20-21^. These many times outweigh the robotic health care expenditure in developing nations^22^.

This case series conducted in developing India presents the first hand scientific evolutionary transition of RC from ORC to LRC specific to neobladder diversion from ileal conduit. An interesting observation during consultation was that Indian patients prefer neobladder over ileal conduit; reason (driving force) for neobladder preference being social acceptability (absence of noticeable urine bag) and preservation of body image. Although more than 55 RC’s were performed, this case series compares neobladder diversion between ORC (n=6) and LRC (n=7). The comparisons emphasize the procedural and outcome variables and the learning experience of the surgeon.

Baseline characteristics were similar between the groups. All patients were young adults, with mean age being 56.5 years with similar existing comorbidities. Also tumor stages were similar in both groups.

Few studies have shown lymph node yield as a marker of surgical quality and accurate staging^23-24^. As compared to other studies reporting a lymphnode yield of 15, in the present analysis the lymph node yield was around 12 in the LRC group, probably attributing to the initial learning curve for laparoscopic cystectomy with the affirmation of improvement^25^. Since the retrieval number was greater than 10, as per National Comprehensive Cancer Network^®^ (NCCN^®^) guidelines, the oncological outcomes were not compromised. To support it with the fact that no PSM were reported in the case series.

In LRC patients, the midline incision length was significantly smaller (P<0.0001) impacting short term hospital outcomes and quality of life parameters. Although the operative time was longer in LRC group as compared to ORC it was sufficed by reduced time for analgesics, and shorter hospital stay besides earlier time to liquid intake with immediate removal of nasogastric tube. This finding is similar to study by Hemal and Kolla (2007)^23^. Blood requirement in terms of transfusion was similar in both groups, this was not surgically inclined by attributed mainly to maintain high Hb content in the patients to speed the healing process post operatively.

Although the operative time was longer in the LRC group, associated complications were relatively low as compared to ORC patients. No major complications were observed in the LRC group unlike ORC group where in one patient died due to sepsis, wound dehiscence and intestinal obstruction. Our study confirmed that LRC with minimal incisions has additional benefits of less pain, less complications, shorter recovery time, without compromising early oncological outcomes.

The RAZOR trial showed non-inferiority of robotic assisted cystectomy vs open, however comparative data with LRC are few^26^. Such case series will help build evidences of neobladder diversions with LRC.

Several limitations of this case series including small sample size with short term follow-up will be addressed with larger sample size and longer follow ups in considerations of the pathophysiological findings.

Based on the observations of our small study sample peri and postoperative outcomes are promising for LRC compared to ORC for patients undergoing neobladder in terms of the smaller incision length associated with less pain and complications, with speedy recovery without jeopardizing oncological outcomes.

## Data Availability

All data produced in the present study are available upon reasonable request to the authors. Data is obtained from Hospital medical records

